# CURATE.AI – AI-derived personalized tacrolimus dosing for pediatric liver transplant: A retrospective study

**DOI:** 10.1101/2022.11.24.22282708

**Authors:** Shi-Bei Tan, Kirthika Senthil Kumar, Tiffany Rui Xuan Gan, Anh T. L. Truong, Lester W. J. Tan, Agata Blasiak, Vidyadhar Padmakar Mali, Marion M. Aw, Dean Ho

## Abstract

Tacrolimus is the cornerstone of immunosuppressive therapy after pediatric liver transplantation. However, reliance on the physician’s experience for dose titration, coupled with tacrolimus’s narrow therapeutic window and inter and intra-patient variability, often results in frequent under or over-dosing with detrimental patient outcomes. Existing predictive dose personalization models are not readily feasible for clinical implementation, as they require multiple measurements each day while the standard frequency is once daily. We developed CURATE.AI, a small-data artificial intelligence-derived platform, as a clinical decision support system to personalize doses using the patient’s own data obtained once a day. Retrospective dose personalization with CURATE.AI on 16 patients’ data demonstrated potential to enable patients to stay in the therapeutic range longer and reach the therapeutic range significantly earlier. Our findings support the testing of CURATE.AI in a prospective controlled trial as an aid for the physician’s decision on tacrolimus dose personalization after pediatric liver transplantation.

## 1. Introduction

Liver transplantation (LT) is an established treatment for children with decompensated liver disease, liver-based metabolic disorders, acute liver failure, and unresectable primary liver malignancy^1^. Continued improvements in surgical techniques, peri-operative care and immunosuppression over the last 40 years have enabled a 5-year survival rate of over 85% in pediatric LT^1,2^. These resulted in the shift of focus to improving the morbidity associated with immunosuppression regimens^1^, particularly in minimizing the risks of acute or chronic liver rejection^1^ and toxicity risks such as nephrotoxicity and neurotoxicity in the long term^3^.

Post-transplant immunosuppression is critical for minimizing the risk of acute or chronic graft rejection^1^, there is morbidity associated with immunosuppression in the long-term; including impact on growth, nephrotoxicity and neurotoxicity ^5^. Tacrolimus is the most common immunosuppressive agent utilised in pediatric LT ^6,7^. The current standard-of-care (SOC) for post-transplant immunosuppression is personalized empirical tacrolimus dose adjustments based on tacrolimus trough levels (TTL). The personalized dose adjustments are guided as per the experience of the transplant physician^4^. Furthermore, tacrolimus has a narrow therapeutic window and has inter- and intra-subject pharmacokinetic variability^5^. As a result, SOC dosing results in frequent deviations from the target therapeutic range ^6^, with the risk of clinically significant adverse effects due to under or over-immunosuppression. With a greater cumulative exposure to immunosuppressive agents that occurs throughout their lifetime, thereby increasing the risk of associated morbidity^7^, there is a dire need for an optimal immunosuppression in pediatric LT.

Most technological advancements for personalized post-transplantation dosing of tacrolimus are for adult liver and kidney transplant patients. Current personalized dosing models for pediatric transplant are based on the area under the curve (AUC) of concentration-time plots which require multiple concentration-time points across the dosing interval i.e. increased frequency of data collection (blood testing) throughout the day when compared to SOC^8,9^, which may not be practical. A study that explored using 13 machine learning models for personalized predictions of TTL for infant liver transplant patients identified that those models required 3 to 7 parameters ranging from graft to recipient weight ratio to specific genotypes and level of albumin in the blood, which can be resource-intensive^10^. As such, for pediatric LT, there remains a need for a solution that is simple, practically applicable with minimal or no additional resources and parameters.

CURATE.AI is a mechanism-independent and indication-agnostic artificial intelligence (AI)-derived dose optimization platform that only uses an individual patient’s data of drug dose and phenotypic treatment response to calibrate a personalized response profile. It identifies then recommends an optimal dose based on the individual’s personalised profile to achieve the target treatment response for the patient. The profile of phenotypic responses across a dose range is represented as a smooth second-order surface. This second-order relationship was previously identified through neural network analysis^11^, and subsequently experimentally validated for various indications^12-14^. Of note, lower order polynomials of the first order or higher order polynomials may also be applicable depending on the intervention and disease. Implementing this platform is markedly simple without the need for neural network analysis and with high clinical actionability. The CURATE.AI platform has already been validated for tacrolimus dose optimization of post-liver transplant immunosuppression to maintain target TTL in adult patients^15,16^. CURATE.AI is currently studied in several clinical trials, including dose optimization trials for tacrolimus in adult liver and kidney transplants and to treat solid and hematological cancers (NCT03527238, NCT04522284, NCT05175235, NCT05381038, NCT04522284, NCT04848935, NCT03759093, NCT04357691). Its usage has also been explored in hypertension management (NCT04769141, NCT05376683) and in tandem with a digital intervention^13,17,18^.

This study is purposed to lay the foundational work for a larger prospective study to determine the impact of personalized tacrolimus dosing in pediatric liver transplant patients. In this retrospective optimization study, CURATE.AI’s applicability in pediatric liver transplant is evaluated first by establishing a linear tacrolimus dose-response relationship for 16 pediatric liver transplant patients. CURATE.AI’s performance was evaluated with both technical and clinically relevant performance metrics. CURATE.AI’s potential clinical actionability was explored by simulating patient journeys and assessing if potential dose recommendations could achieve the therapeutic range earlier and increase the time in the therapeutic range.

## 2. Methods

### 2.1 Study design and data collection

This retrospective study analyzed de-identified data of 16 pediatric liver transplant patients that were collected from existing medical records at the National University Hospital in Singapore, the only pediatric liver transplant centre in Singapore. Data were obtained during the period from 1 January 2011 to 31 December 2020, as approved by the Centralized Institutional Review Board (CIRB reference: 2019/2040). Inclusion criteria were pediatric (between ages 1 – 16) living donor, liver transplant patients on their Index admission for transplant. Exclusion criteria were: re-transplant cases, cases with mortality, and cases with complex clinical scenarios that required re-exploration within the Index admission because of the multiple confounders.

### 2.2 Data processing

CURATE.AI required two components to be supplied into the platform as inputs: the administered tacrolimus dose in mg (dose), and the corresponding TTL in ng/ml (response), to curate a personalized profile for each patient. While the used records showed the doses had been administered twice a day (BID), the dose input for CURATE.AI was the effective 24-hour dose and the corresponding TTL response measured the next day, right before the morning dose. Days with two measurements of TTL (taken in the morning and evening) instead of one, which is the standard frequency, were excluded. Further, measurement equipment was unable to provide resolution for TTL less than 2 ng/ml. Hence, TTL measurements < 2 ng/ml were assessed to be inaccurate and thus excluded. Doses recorded from the day of discharge onwards were assessed to be unreliable, as confirmation of the doses administered was not possible, and thus excluded. This study used data points, also known as dose-response pairs, over the set of consecutive days for each patient for the analysis by CURATE.AI. Dose ranges were categorized as low (< 2 mg), medium (2 mg to < 4 mg), and high (> 4 mg).

### 2.3 CURATE.AI

The flow of the analysis with CURATE.AI is illustrated in **Fig. 1**. Linear regression was used to model the relationship between the tacrolimus dose and the corresponding TTL (response) and forms the individualized profile of the patient. Only 2 dose-response pairs were required for the linear regression to provide an individualized profile. Where the profile intersected with the therapeutic range, defined as between and inclusive of 8 and 10 ng/ml for every patient, doses in multiples of 0.5 mg (the smallest tacrolimus capsule available^19^) were identified, representing the potential doses that the patient might have benefitted from with CURATE.AI-assisted dosing. When there are multiple possible doses that fulfil the requirements, the lowest dose was considered as the CURATE.AI recommendation. The profile, which consisted of 2 dose-response pairs from 2 dosing events, was used to predict the TTL of the next dosing event. The difference between the observed TTL and the predicted TTL was defined as the prediction error.

**Fig. 1:**
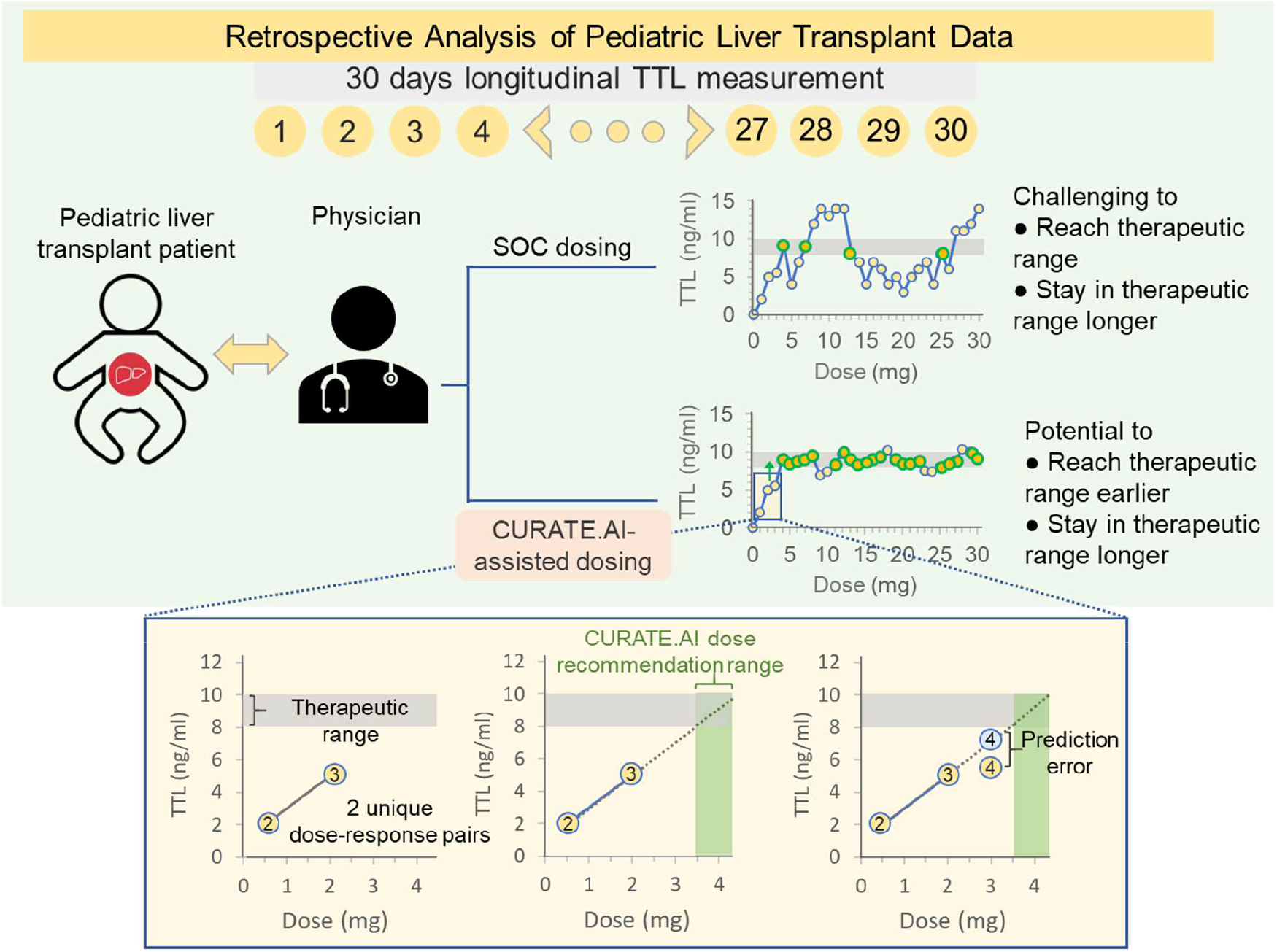
Retrospective analysis of pediatric liver transplant data. Data from pediatric liver transplant patients were collected for the duration of 30 days post-transplant and were retrospectively analyzed. SOC dosing relies on the unaided decisions by the physicians, with challenges to reaching the therapeutic range fast and staying within it. CURATE.AI-assisted dosing aims to enable the TTL to reach the therapeutic range earlier and stay in the therapeutic range longer. The enlarged section below depicts an example of a dose-response relationship, also known as the individualized profile, based on the 2^nd^ and 3^rd^ day post-transplant. CURATE.AI-assisted dose recommendation for the 4^th^ day is based on the intersection between the profile and the therapeutic range. A prediction of the TTL after the next dosing event was made based on the patient’s CURATE.AI profile, and the prediction error was defined as the difference between the observed and predicted TTL.

Only 2 dose-response pairs were used as a calibration to provide a dose recommendation for the next dosing event. The choice of the 2 pairs requires using a rolling window approach, defined as using a window of a specified size of 2 that rolls through the data, 1 unit a time for each profile generation. This means that dose recommendation can only start after 2 dose-response pairs are obtained, requiring at least 2 days with unique doses and the corresponding TTL.

### 2.4 Performance Metrics

CURATE.AI’s performance was evaluated based on technical performance metrics widely used in literature to evaluate predictive models. Additionally, clinically relevant performance metrics were devised to characterize the potential clinical actionability of CURATE.AI as a CDSS.

The technical performance metrics were based on prediction errors, defined as the differences between the observed TTL and the TTL value predicted by CURATE.AI. Specifically, the technical performance metrics used were prediction error, absolute prediction error, and root mean squared error (RMSE). CURATE.AI’s absolute prediction error was compared to the prediction errors from the machine learning models by Song et al^10^, as it also predicted TTLs.

For clinically relevant performance metrics, contextual information on pediatric liver transplant immunosuppression were considered in devising the metrics. Specifically, the clinically acceptable prediction error was defined as between and inclusive of -1.5 and 2 ng/ml, based on the clinically acceptable range of TTL of 6.5 to 12 ng/ml, and in consideration of the therapeutic range of between and inclusive of 8 and 10 ng/ml (**Supplementary Material Fig. S2**). The percentage of predictions within clinically acceptable prediction error was computed. The predictions outside of the clinically acceptable prediction error were categorized as overpredictions and underpredictions, defined as less than -1.5 ng/ml and greater than 2 ng/ml, respectively. The percentages of overpredictions and underpredictions were computed. The numbers of predictions that satisfy the following 5 assessment criteria were computed in steps illustrated in a flow chart (**Fig. 4**). The first 2 criteria focused on CURATE.AI’s prediction of TTL with the administered doses, and the next 3 criteria focused on CURATE.AI’s dose recommendations’ ability to achieve the therapeutic range. The criteria were: 1) CURATE.AI profile reliability (considered reliable if CURATE.AI correctly predicted that TTL would fall within the therapeutic range, pre-defined as between and inclusive of 8 and 10 ng/ml, or non-therapeutic range), 2) CURATE.AI prediction accuracy (considered accurate if CURATE.AI predictions were within the range of clinically acceptable prediction error, pre-defined as between and inclusive of -1.5 and 2 ng/ml), 3) whether the CURATE.AI-recommended dose differed from the dose administered, 4) whether the observed TTL was outside of the therapeutic range, and 5) dose actionability of the CURATE.AI-recommended dose (considered actionable if CURATE.AI dose recommendations were 8 mg or below).

2 representative patient cases were investigated to identify potential scenarios in which using CURATE.AI could be beneficial. The projected effects of utilizing CURATE.AI on each day of the treatment were explored, and the effects of CURATE.AI were categorized into 3 groups, namely: ‘no effect’, ‘improve’, or ‘worsen’ the time in therapeutic range. The effects are based on the 5 assessment criteria listed previously (**Supplementary Material Fig. S1)**. The projected percentage of days within the therapeutic range, the number of days required to first achieve the therapeutic range, and the percentage of patients that could achieve the therapeutic range in the first week were computed for the modelled scenarios with CURATE.AI-assisted dosing and compared with SOC dosing.

## Statistical analysis

The normality of the data distribution was tested with the Shapiro-Wilk test. Continuous variables were represented as mean ± standard deviation (S.D.) or median (interquartile range (IQR)), depending on the normality of the data type. Wilcoxon signed-rank test was used to compare the medians of two non-parametric groups. Bartlett’s test was used to check for equal variance between two unrelated parametric groups. An unpaired t-test was used to compare the means of two unrelated parametric groups with equal variance. Paired t-test was used to compare the means of two related parametric groups. The statistical significance was defined as *p* < 0.05.

## 3. Result

### 3.1 Patient population

The characteristics of the 16 pediatric liver transplant patients whose de-identified data were included in this study are listed in **Table 1**.

**Table 1.** Patient characteristics.

| Characteristics, Units | Pediatric liver transplant patients (N = 16) |
| --- | --- |
| Gender | Male, 13 |
|  | Female, 3 |
| Age, years | 2 (0.98 – 8.25)* |
| Body mass index, kg/m<sup>2</sup> | 17.45 (16.03 – 18.55)* |
| Ethnicity | Chinese, 15 |
|  | Malay, 1 |
| Number of days with blood draw results | 20 (13 – 22)* |
| Child-Pugh score | B8 (A6 – C10)* |

\* Median (IQR). Values for the Child-Pugh Score are calculated based on the numeric.

Inter-individual heterogeneity was observed in the longitudinal data **(Fig. 2)**. Out of all 16 patients, 15 (93.75%) patients achieved the therapeutic range at least for 1 day. Patients who achieved the therapeutic range did so at varied dose ranges. Out of the 15 patients that achieved the therapeutic range at least for 1 day, 2 (13.33%) patients achieved the therapeutic range at low doses only, 3 (20.00%) patients at medium doses only, 1 (6.67%) patient at high doses only, 4 (26.67%) patients at both low and medium doses, 3 (20.00%) patients at both medium and high doses, and 1 (6.67%) patient across all dose ranges. The patients deviated from the therapeutic range most of the time and stayed within the therapeutic range for a mean of 23.94 ± 15.29 % of days. The patients first achieved the therapeutic range within a median of 7.00 (IQR 4.50 – 8.00) days. Out of the 15 patients that achieved therapeutic range for at least 1 day, 10 (66.67%) patients achieved the therapeutic range within the first week. The mean TTL was 8.79 ± 3.26 ng/ml (n = 287 TTL) and the mean dose was 2.63 ± 1.49 mg (n = 276 doses), corresponding to a mean of 0.21 ± 0.16 mg/kg/day.

**Fig. 2:**
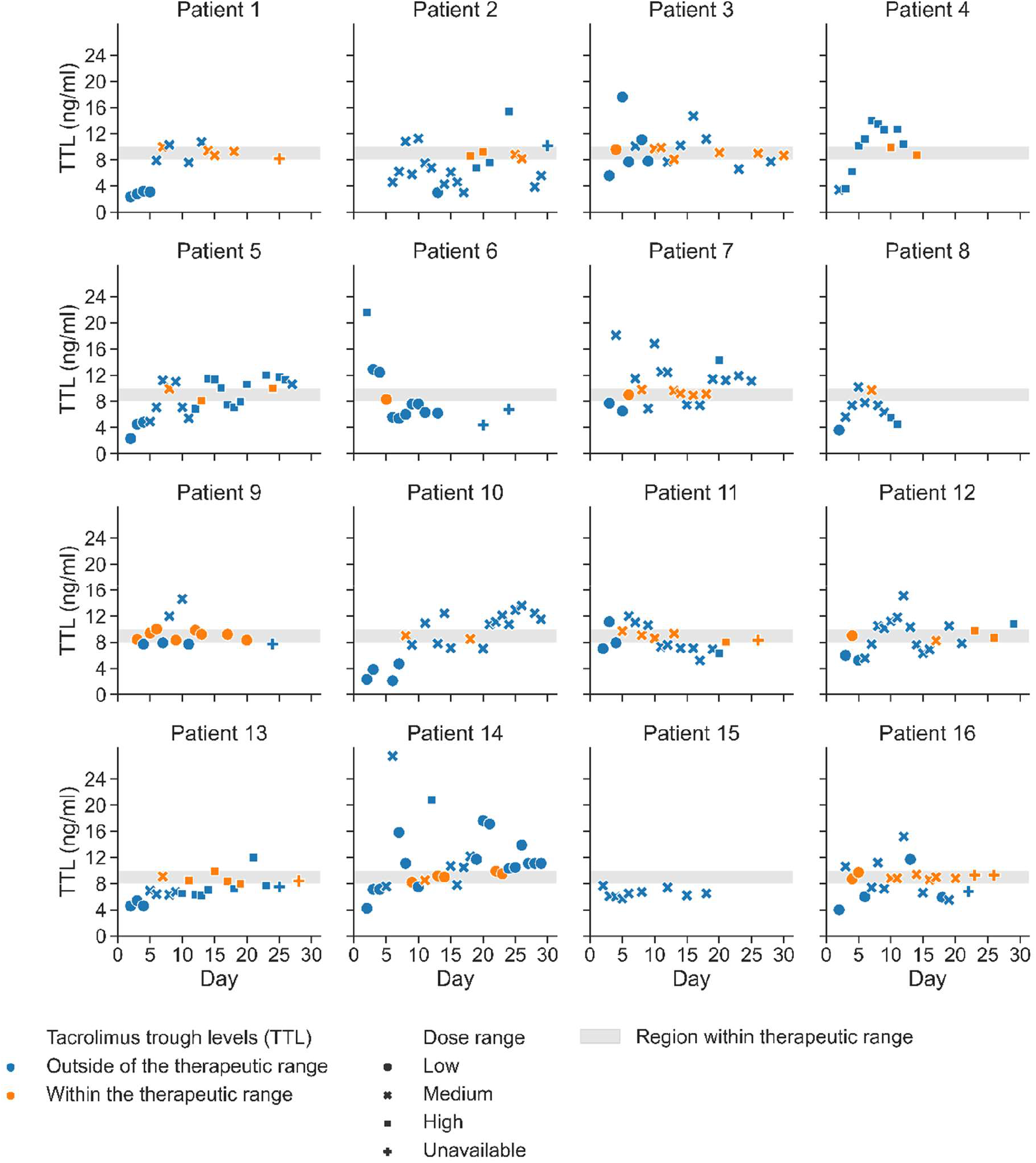
TTL over time of each patient. The grey region represents the therapeutic range of between and inclusive of 8 and 10 ng/ml. Blue and orange markers represent TTL outside and within the therapeutic range, respectively. Dose ranges are categorized as low (less than 2 mg, circle markers), medium (2 to less than 4 mg, square markers), and high (4 mg and higher doses, cross markers), respectively. Unavailable doses refer to missing or unreliable (from the day of discharge onwards) doses, and data points without TTL are not reflected here. Inter-individual heterogeneity was observed in terms of the dose ranges in which patients achieved the therapeutic range, the duration that patients stayed within the therapeutic range, and the number of days taken to first achieve the therapeutic range.

The flow of data inclusion for CURATE.AI analysis is illustrated in **Fig. 3**. The data of all 16 patients passed the first stage screening requirement of minimally 2 uniquely modulated doses and available corresponding TTL. 3 patients’ data were excluded in the second stage of data screening due to too few predictions, defined as less than 3 predictions. 13 patients met the criteria of the 2 screening stages and were included for subsequent retrospective CURATE.AI applications and analysis up to **Section 3.4**.

**Fig. 3:**
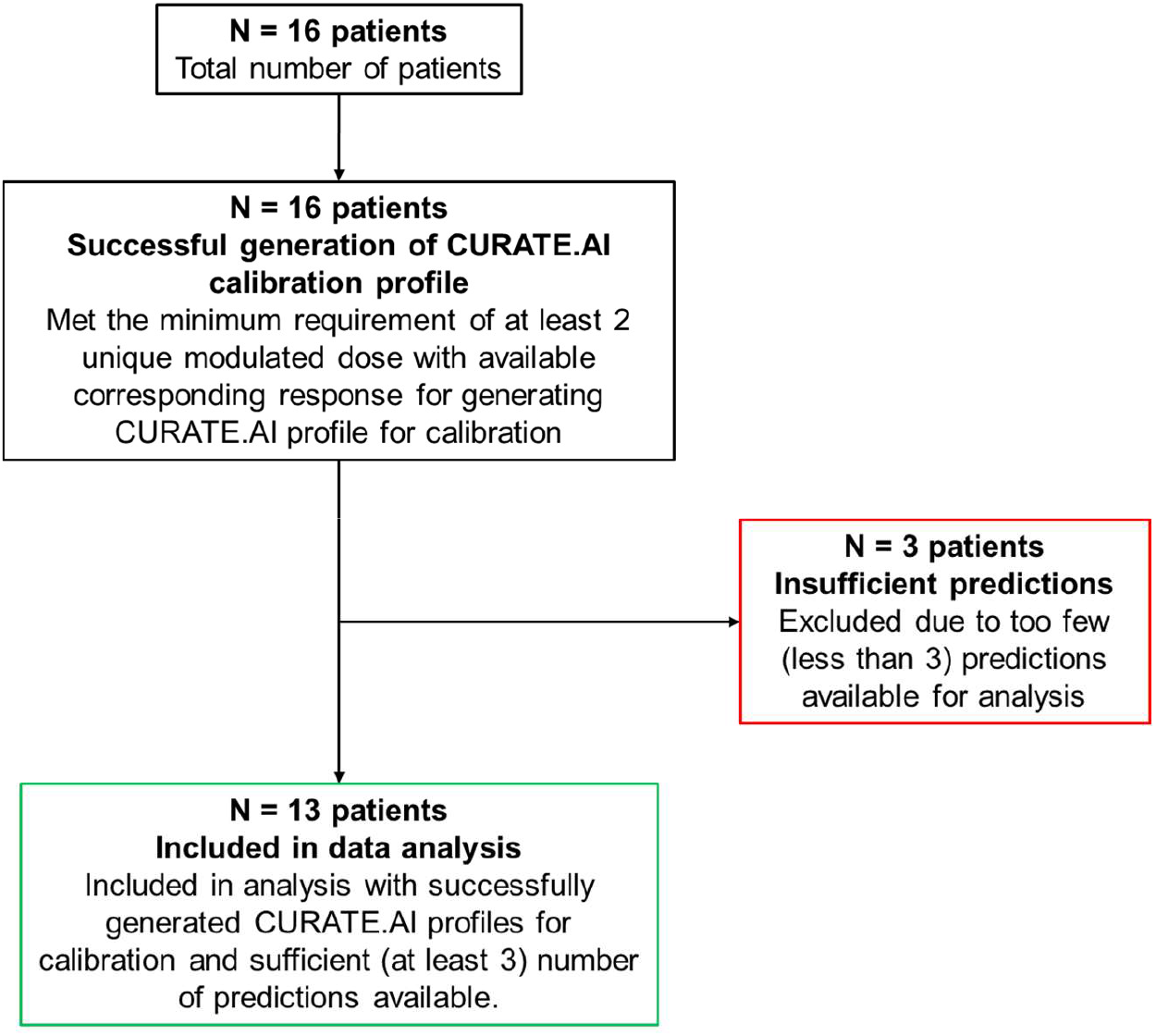
Flow of patient data inclusion for CURATE.AI analysis.

### 3.2 Evaluation of CURATE.AI performance

#### 3.2.1 Technical performance

CURATE.AI demonstrated satisfactory predictive performance with a low mean prediction error of 0.19 ± 2.75 ng/ml (n = 121 predictions) and an acceptable median absolute prediction error of 1.80 (IQR 0.80 – 2.80) ng/ml which is comparable to the mean absolute prediction error (mean: 2.01 ng/ml, mean + S.D.: 3.35 ng/ml, mean - S.D.: 0.85 ng/ml) of the best-performing machine learning model by Song et al that predict TTLs for infant liver transplant patients. The RMSE was 2.75 ng/ml for all predictions (n = 121 predictions).

#### 3.2.2 Clinically relevant performance

CURATE.AI achieved 47.11 % (n = 121) of the predictions within the pre-defined clinically acceptable prediction error, pre-defined as between -1.5 and +2 ng/ml. Overpredictions and underpredictions that exceeded the clinically acceptable prediction error, pre-defined as less than -1.5 ng/ml and greater than 2 ng/ml respectively, comprised 28.10 % and 24.79 % (n = 121) of the predictions respectively.

All CURATE.AI predictions were further assessed for the potential of improving patients’ responses (**Fig. 4)**. An actionable and accurate CURATE.AI profile was generated for 37.19% (n = 121) of the days, indicating that CURATE.AI had the potential to identify doses for those dosing events to achieve the therapeutic range. The use of CURATE.AI to augment dosing decisions, in turn, could lead to achieving a similar or higher percentage of days within the therapeutic range, as compared to the observed TTL in the data collected.

**Fig. 4:**
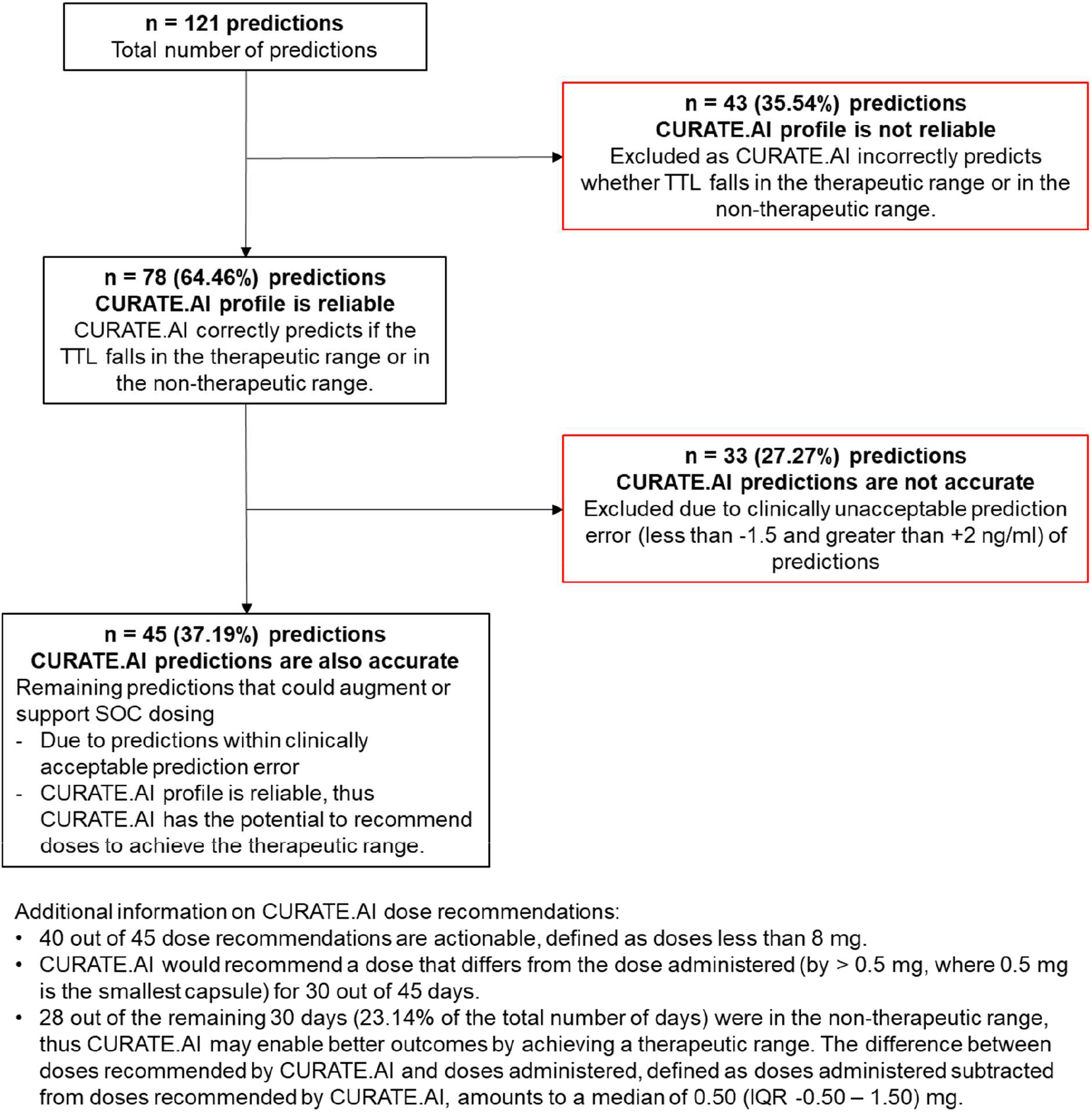
Assessment of predictions for augmenting or supporting dosing.

Out of the days in which the reliable and accurate CURATE.AI profiles were generated, the CURATE.AI dose recommendations were further assessed for dose actionability (pre-defined as a maximum of 8 mg) of the recommended dose, whether the recommended dose differed from the dose administered, and whether the observed TTL was within the non-therapeutic range. CURATE.AI-assisted dosing could have potentially achieved the therapeutic range on 23.14% of the days (n = 121). Additionally, the difference between the doses recommended by CURATE.AI and the doses administered, defined as the doses administered subtracted from doses recommended by CURATE.AI, amounted to a median of 0.50 (IQR -0.50 – 1.50) mg.

### 3.3 Case series

We investigated 2 representative patient’s care which followed different dosing strategies to demonstrate clinical use cases where CURATE.AI may be beneficial.

### Case 1: CURATE.AI may lead to achieving the therapeutic range earlier

Patient 5 received a wide range of tacrolimus doses from 0.5 to 5.0 mg. Patient 5 achieved the therapeutic range only on Days 8, 13, and 24 (**Fig 5a**), out of the 27 days with available data.

**Fig. 5:**
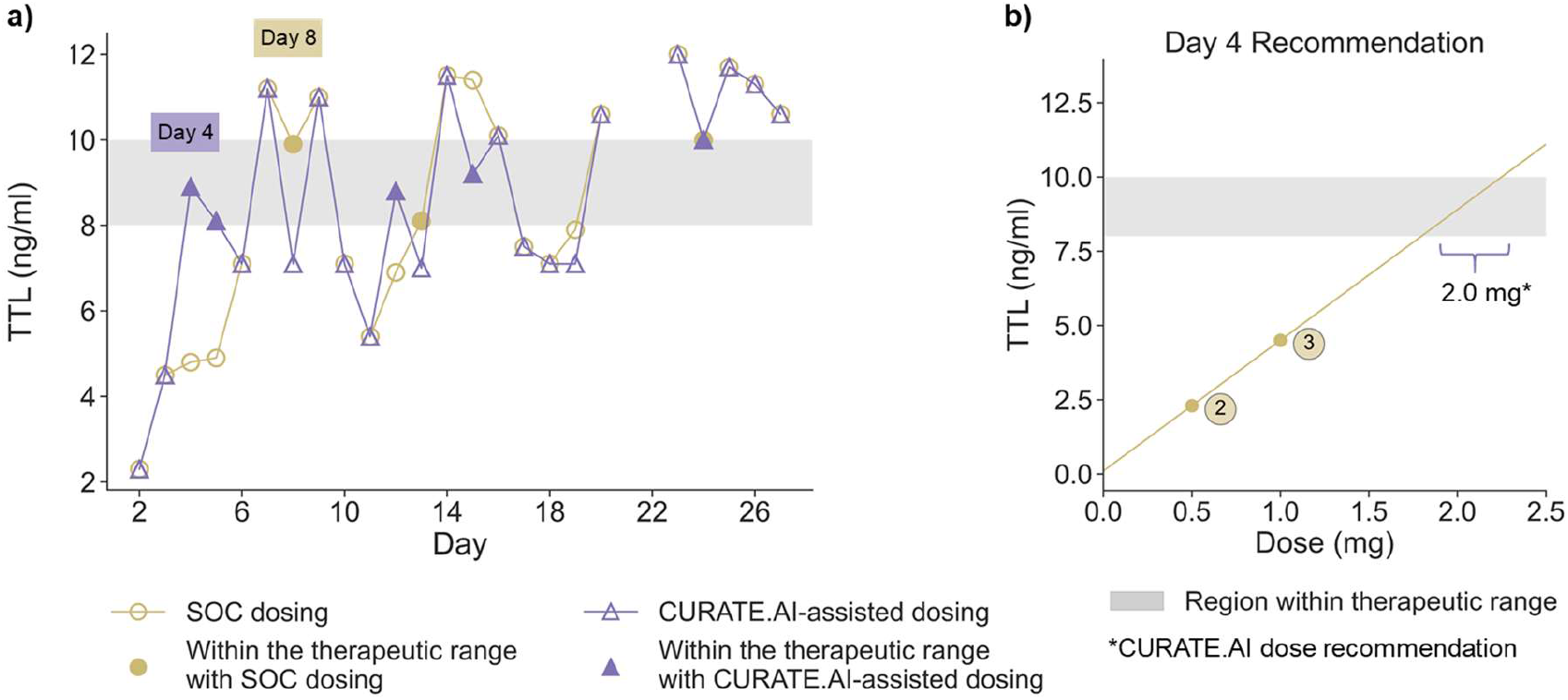
Patient 5’s treatment journey with and without CURATE. **AI**. The grey region in both graphs represents the region within the therapeutic range, defined as between and inclusive of 8 and 10 ng/ml. **a** TTL achieved during SOC dosing (yellow circle markers without fill), TTL within the therapeutic range during SOC dosing (yellow circle markers with fill), projected TTL with CURATE.AI-assisted dosing (purple triangle markers without fill), projected TTL within the therapeutic range with CURATE.AI-assisted dosing (purple triangle markers with fill) are indicated. TTL on Days 21 and 22 were missing from the data collected. **b** Doses given on Days 1 and 2, leading to TTL on Days 2 and 3, are depicted in yellow circle markers without fill. The yellow points are numbered by the days on which the corresponding TTL was observed. CURATE.AI’s dose recommendation of 2.0 mg would potentially enable achieving the therapeutic range on Day 4.

For this patient, CURATE.AI successfully generated a profile from the 2 dose-response pairs from Days 2 and 3 (**Fig. 5b**). Based on the generated profile, CURATE.AI would have recommended 2.0 mg of tacrolimus (instead of the 1.5 mg dose administered) (**Fig 5b**) which had the potential to lead to achieving the therapeutic range 4 days earlier, on Day 4 instead of Day 8 (**Fig. 5a**). Following that first CURATE.AI-guided dosing event, the new data pair would have been incorporated back into the profile to ensure the profile evolves with the patient state and new dose recommendations can be generated over time. Based on the fact that a reliable CURATE.AI profile was obtained for 15 out of next 23 days with complete data for this patient, and CURATE.AI suggested different doses than the administered doses on 10 of the days, it is plausible that using CURATE.AI would not only have led the TTL to achieving the therapeutic range faster but also a higher percentage of days when the therapeutic range was achieved.

CURATE.AI was able to generate actionable profiles and dose recommendations for 15 days out of the next 23 days where doses were administered, where CURATE.AI suggested different doses than the administered doses on 10 of the days.

### Case 2: CURATE.AI’s dynamic adjustment may enable sustained TTL within the therapeutic range

Patient 4 received 3 mg of tacrolimus on Day 1, which led to TTL outside of the therapeutic range on Day 2. Subsequently, the patient received 4 mg on Day 2, which again led to TTL outside of the therapeutic range on Day 3. Afterwards, the patient received 6 mg for 11 consecutive days. The patient’s TTL only achieved the therapeutic range on Day 10, after 7 consecutive days of 6 mg, and fell out of the therapeutic range on Days 11 and 12 when 6 mg was administered (TTL was unavailable on Day 13 when 6 mg was administered), and achieved the therapeutic range again on Day 14 when 6 mg was administered.

For this patient, CURATE.AI successfully generated profiles for dose recommendations for Day 3 onwards. **Fig. 6a** illustrated the profiles generated for dose recommendations from Days 5 to 9 based on dose inputs of 4 mg on Day 3 and the day before the day of recommendation and the corresponding TTL. The slopes of the profiles varied in steepness over time with the same dose inputs of 4 mg and 6 mg, suggesting longitudinal variability in TTL response to tacrolimus dose. The dynamic TTL responses from the static and repeated doses over consecutive days suggest that the patient’s state was changing, and repeatedly administering the same dose over consecutive days was not an optimal dosing strategy. With dose titrations recommended by CURATE.AI (**Fig. 6b**), the patient’s TTL plausibly could have achieved the therapeutic range more often, and potentially sustain being in the therapeutic range more often throughout the treatment (**Fig. 6a**). The projected TTLs, defined as the estimated TTLs if CURATE.AI’s actionable dose recommendations were administered, ranged from 8.55 to 14.00 ng/ml with CURATE.AI-assisted dosing for Days 5 to 9, compared to the observed TTLs of 11.2 to 14.00 ng/ml which were all outside of the therapeutic range (**Fig. 6c**).

**Fig. 6:**
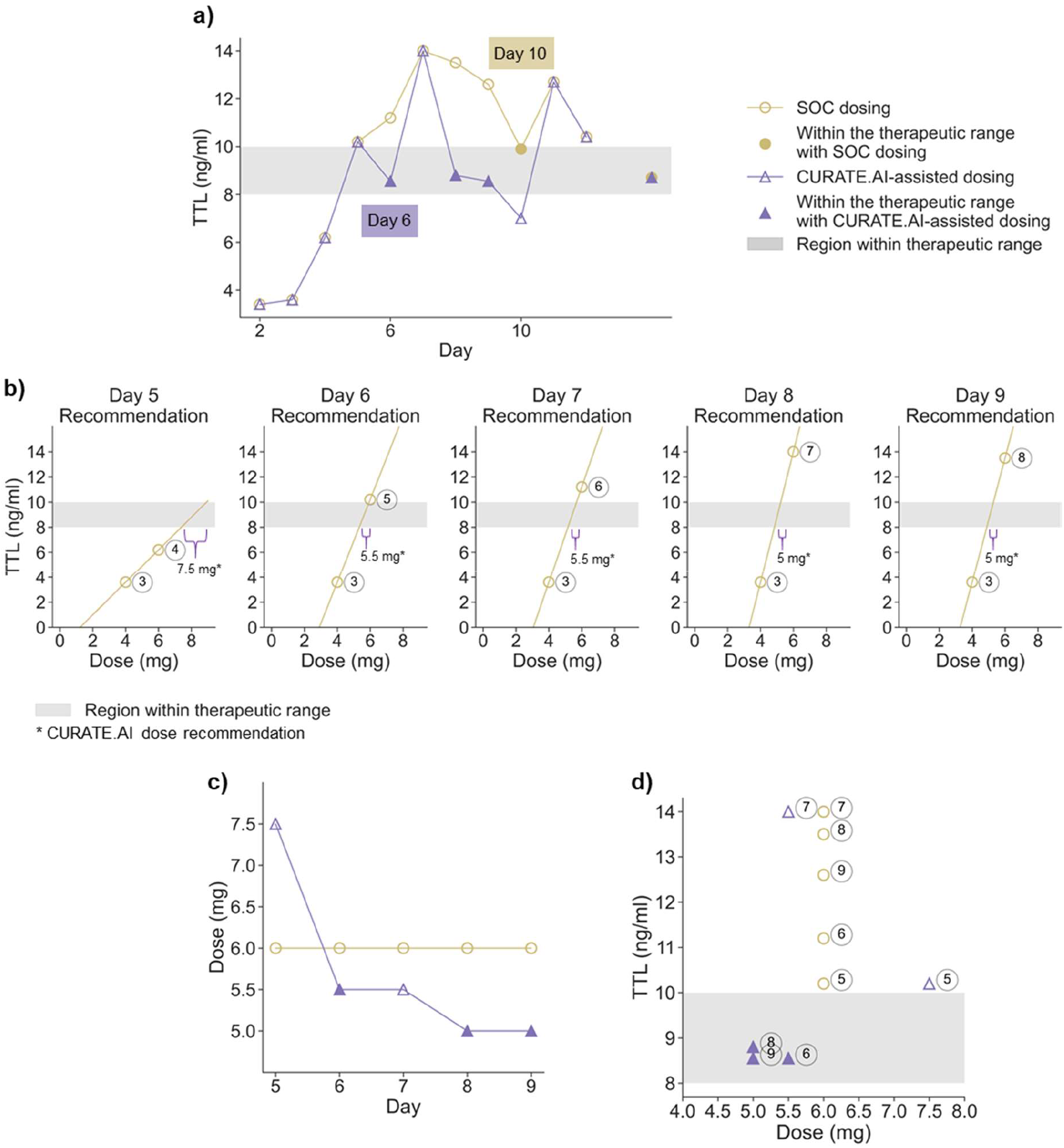
Patient 4, CURATE.AI dose modulations to achieve and sustain the therapeutic range. **a** TTL achieved during SOC dosing (yellow circle markers without fill), TTL within the therapeutic range during SOC dosing (yellow circle markers with fill), projected TTL with CURATE.AI-assisted dosing (purple triangle markers without fill), projected TTL within the therapeutic range with CURATE.AI-assisted dosing (purple triangle markers with fill) are indicated. TTL on Day 13 was missing from the data collected. **b** profiles generated for dose recommendations from Days 5 to 9 based on dose inputs of 4 mg on Day 3 and the day before the day of recommendation and the corresponding TTL profiles generated for dose recommendations from Days 5 to 9 based on dose inputs of 4 mg on Day 3 and the day before the day of recommendation and the corresponding TTL. Dose recommendations are indicated within the range of the purple brace. The actionable doses recommended (in multiples of 0.5 mg) are indicated under the purple brace. **c** Doses administered over Days 5 to 9 (yellow circle markers) and the recommended doses by CURATE.AI (purple triangle markers) are indicated. **d** TTL corresponding to Days 5 to 9 (yellow circle markers) and the projected TTL with CURATE.AI dose recommendations (purple triangle markers) are indicated. The grey region spans the therapeutic range.

### 3.4 CURATE.AI’s effect and inter-individual differences

Retrospective dose optimization was simulated using the data of all 16 patients (**Fig. 7a**). Out of a total of 276 days across data from all 16 patients, the observed TTL moved from outside to within the therapeutic range for 10.14% of the days; the observed TTL in the therapeutic range fell out of the therapeutic range for 5.43% of the days; and the observed TTL stayed within or outside the therapeutic range for 84.42% of the days. The criteria for determining the effect of CURATE.AI can be found in **Supplementary Material Fig. S1**. Notably, the simulation results demonstrated improvement in the patients’ treatment responses across the whole timespan of the treatment.

**Fig. 7:**
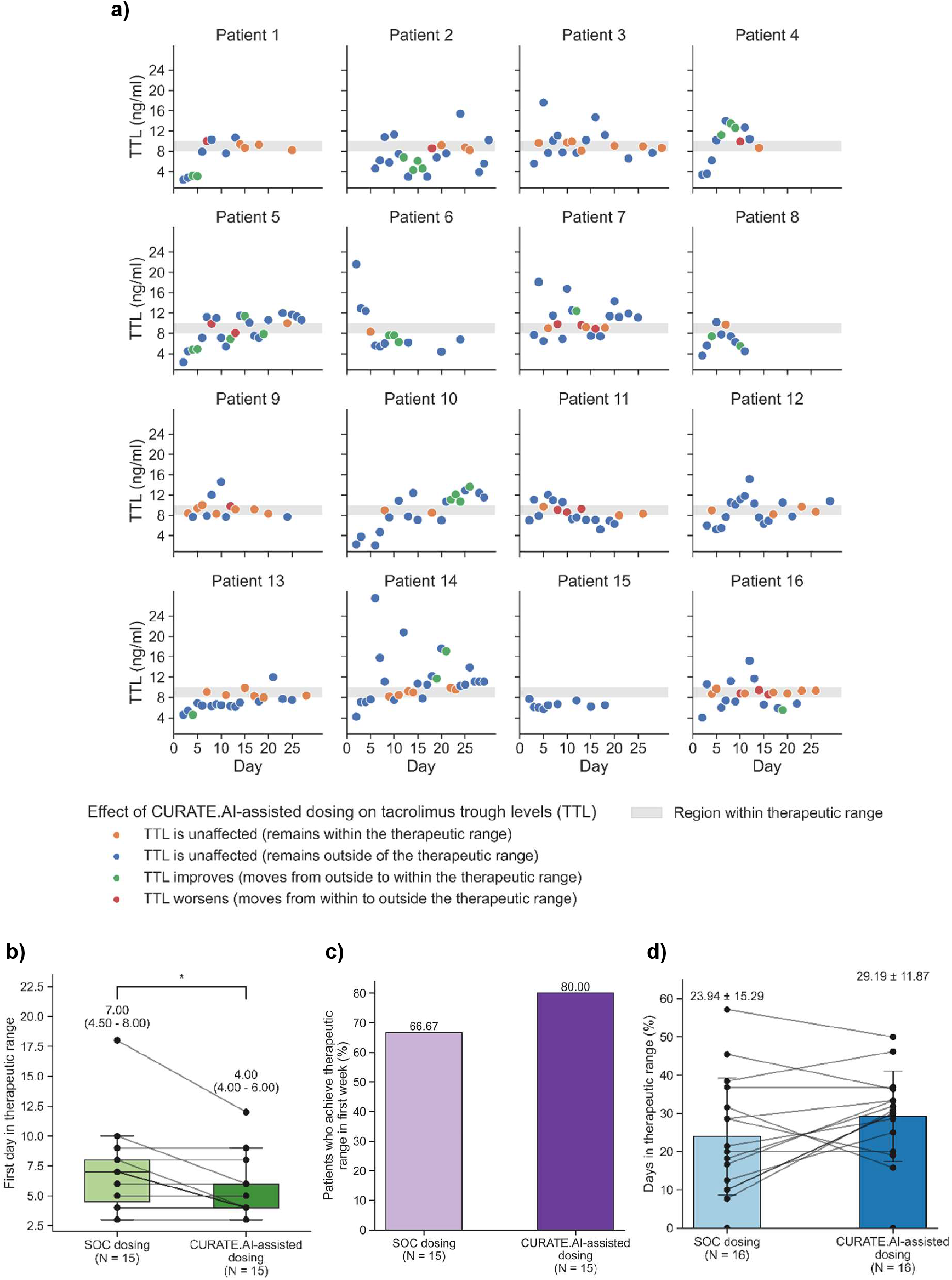
Projected effects of CURATE.AI. **a** Results of the retrospective dose optimization simulation with CURATE.AI-assisted dosing is presented. The grey region represents the therapeutic range. Blue and orange circle markers represent TTL that are projected to remain within and outside of the therapeutic range, respectively. Green and red markers represent TTL that are projected to move from outside to within the therapeutic range and vice versa, respectively. Dose ranges are categorized as low (< 2 mg, circle markers), medium (2 to < 4 mg, square markers), high (> 4 mg, cross markers), and unavailable (plus markers), respectively. Data points without either TTL or dose are not reflected here. The results of SOC dosing and projected results with CURATE.AI-assisted dosing are illustrated in **b – d**, where each scatter point represents data from a single patient, and lines linking 2 scatter points across bars or boxplots represent linkage between the same patient. **b** First day in the therapeutic range (boxplot represents the minima and maxima, lower and upper quartiles, and the medians, N = 15 patients). * indicates a statistically significant difference with *p* = 0.03 < 0.05 estimated with Wilcoxon signed-rank test, between the first day in the therapeutic range as observed during SOC dosing and predicted with CURATE.AI-assisted dosing. **c** Percentage of patients (N = 15 patients) who achieved the therapeutic range in the first week. **d** Percentage of days in therapeutic range (bar plot shows the mean and standard deviation, N = 16 patients).

Furthermore, CURATE.AI had the potential to improve the patient outcomes **(Fig. 7b-d)**. Out of all 16 patients, 6 (37.5%) patients might have achieved the therapeutic range earlier with CURATE.AI-assisted dosing and no change on the rest. The projected median number of days for each patient’s TTL to reach the therapeutic range with CURATE.AI-assisted dosing was 4.00 (IQR 4.00 – 6.00) days (N = 15 patients), which was significantly earlier (*p* = 0.03) than compared to the effects of SOC dosing (7 days, IQR 4.50 – 8.00 days, N = 15 patients) (**Fig. 7b**). Out of the 15 patients that achieved the therapeutic range for at least 1 day, 12 (80%) patients’ TTL reached the therapeutic range within the first week with CURATE.AI-assisted dosing, which is more than recorded after SOC dosing (66.67%) (**Fig. 7c**).

Out of all the 16 patients, CURATE.AI-assisted dosing was projected to have varied effects across patients. CURATE.AI-assisted dosing might enable 9 (56.25%) patients to achieve therapeutic range more frequently, 3 (18.75%) patients equally frequent, and 4 (25.00%) patients less frequently. The mean projected percentage of days within the therapeutic range with CURATE.AI-assisted dosing was 29.19 ± 11.87 % (N = 16 patients) similar (*p* = 0.10) to SOC dosing of 23.94 ± 15.29 % (N = 16 patients) (**Fig. 7d**).

## 4. Discussion

This retrospective study successfully generated individualized profiles for 13 out of 16 children using their individual data consisting of the recorded tacrolimus doses and the corresponding TTL over 30 days following LT. CURATE.AI demonstrated satisfactory performance with both technical and clinically relevant performance metrics.

Other personalized tacrolimus dosing methods proposed for pediatric liver transplant are AUC-based methods^1,2^ that require resource-intensive high-frequency measurements across the dosing interval and face the resource barrier for translation into clinical practice. CURATE.AI is less resource-intensive than any of the 13 machine learning models compared in the recently published study by Song et al^10^ for personalized dosing for infants with liver transplants, as CURATE.AI requires only 2 parameters while the machine learning models required 3 to 7 covariates. Despite using a fraction of inputs, CURATE.AI achieved comparable technical performance – absolute prediction error was comparable to that of the best-performing model described in the study (CURATE.AI’s median absolute prediction error of 1.80ng/ml compared to the mean absolute prediction error of 2.01ng/ml).

Furthermore, CURATE.AI has the potential to overcome the issues of heterogeneity observed in the patient data. Specifically, based on the patient data, the patients achieved the therapeutic range at different dose ranges and/or responded differently over time to the same dose (**Fig. 5 and 6**). The percentage of days within the therapeutic range and the number of days to achieve the therapeutic range also differed across patients. In response to such challenges, CURATE.AI had the potential to identify the optimal doses earlier than SOC, and subsequently adjust the dosing profiles over time to provide dynamic dose recommendations to sustain the patients’ TTL within the therapeutic range over time (**Fig. 7**).

These results suggested that CURATE.AI might be suitable for assisting tacrolimus dosing decisions in pediatric liver recipients to enable similar or better TTL which may translate to improvement in outcomes. However, it is worth noting that CURATE.AI achieved varied results in different patients. Thus, future study is necessary with a larger study sample to identify individuals that are the most likely to benefit from CURATE.AI-assisted dosing.

### 4.1 CURATE.AI in clinical workflow

In current clinical practice, TTL is measured daily; which is aligned with the intended workflow of CURATE.AI to use daily acquired dose-response pairs to generate a profile rapidly. CURATE.AI requires the unique doses and corresponding TTLs over only 2 days to calibrate a personalized profile required to progress to the next step and generate the optimal dose recommendations. A rapidly generated personalized dosing profile would facilitate achievement of optimal TTL within the first week following LT

This study also took into consideration the clinical workflow of dosing modulation by physicians. The initial dose is often based on body weight, and subsequent doses are in increments of 0.5 mg based on the minimum capsule size^19^. Thus, the evaluation of CURATE.AI was based on dose recommendations in increments of 0.5 mg which would ensure the clinical actionability of CURATE.AI’s dose recommendations.

CURATE.AI may facilitate dosing in both short-term and long-term management of tacrolimus in pediatric LT. CURATE.AI is adaptable to changing target therapeutic ranges over the lifespan of the patient, starting from the targeted 8 to 10 ng/ml within 30 days post-transplant, to modified therapeutic ranges in the long term. This will be useful in avoiding graft loss due to immunosuppression-associated complications which will be particularly desirable in the case of Singapore, a country with a relatively small population, shortage of deceased donor organs, and reliance on living donors for pediatric LT ^20^.

### 4.2 Limitations and future directions

The study demonstrated that CURATE.AI may be useful in facilitating prospective management of personalized tacrolimus dosing in pediatric liver transplant patients. However, there are several limitations in this study. The patient data used for the CURATE.AI retrospective optimization are subsets of the patient data, which were the available data over the longest number of consecutive days, and data with only one TTL measurement a day. These choices were made to analyse data that were considered ideal to minimise variability. Also, the TTL measurements and tacrolimus dose administered were not strictly regularly spaced due to practical reasons. Moreover, this retrospective study was conducted with data that were not influenced by CURATE.AI prospectively. With the assistance of CURATE.AI, the initial doses would be varied to enable calibration of a wide range of doses to generate the individualized profile and adopting CURATE.AI dose recommendations over the whole treatment may result in different sets of doses and responses as compared to the static retrospective data evaluated for the effect of CURATE.AI. Thus, the actual effect of CURATE.AI could vary from the projected effect with retrospective data as analysed in this study. Lastly, the sample size of 16 patients is considered small for application to clinical practice. may not be sufficient to conclude that CURATE.AI may be effective in prospective management. However, this study aimed to demonstrate CURATE.AI’s applicability in personalizing tacrolimus dosing, thus the sample size of 16 patients was deemed sufficient for this study. We investigated cases with extreme prediction errors but were unable to provide definitive conclusions on the facts that resulted in the extreme prediction errors, due to the small sample size of the study, and no particular trends were observed.

The results from this study including the satisfactory predictive performance of CURATE.AI and CURATE.AI’s clinical applicability for personalized tacrolimus dosing in pediatric liver transplant patients support further prospective validation.

The advantages of a prospective trial include allowance for additional measurements of TTL with corresponding bespoke dose recommendations by CURATE.AI, a larger recruitment that would enable identification of subgroups characteristics that would or would not benefit from CURATE.AI, and identification of potential factors that lead to extreme prediction errors.

## 5. Conclusion

Appropriate tacrolimus management to achieve and sustain TTLs within the therapeutic range is crucial to avoid liver rejection, nephrotoxicity, neurotoxicity, and other adverse events^1,3^. CURATE.AI is an AI-derived, small data platform, that is mechanism-independent and disease-agnostic. We have demonstrated a proof-of-concept retrospective study of using CURATE.AI to support tacrolimus management through personalized dosing, with promising performance in both technical and clinically relevant metrics. Insights from comparing projected outcomes of CURATE.AI-assisted and SOC dosing highlight the potential of TTL reaching the therapeutic range earlier, sustaining TTL within the therapeutic range longer, and increasing the TTL duration in the therapeutic range. These results set a foundation for a consideration of using CURATE.AI to support the prospective management of tacrolimus dosing in pediatric liver transplants. Future work will focus on testing CURATE.AI in a controlled trial to understand CURATE.AI’s performance using technical and clinically relevant performance metrics and explore the integration of CURATE.AI into the clinical workflow for prospective management of tacrolimus in pediatric liver transplantation.

## Data Availability

The data used in this study are available from the corresponding authors upon reasonable request.

## Acknowledgments

D.H. gratefully acknowledges the National Research Foundation Singapore under its AI Singapore Programme (Award Number:AISG-GC-2019-002), Singapore Ministry of Health’s National Medical Research Council under its Open Fund-Large Collaborative Grant (“OF-LCG”) (MOH-OFLCG18May-0028), and the Ministry of Education Tier 1 FRC Grant.

## Ethics declarations

A.B. and D.H. are co-inventors or previously filed pending patents on artificial intelligence-based therapy development. E.K.-H.C., and D.H. are shareholders of KYAN Therapeutics, which has licensed intellectual property pertaining to AI-based oncology drug development. The findings from this study are being made available for public benefit, and no intellectual property rights arising from the work reported here are being pursued. D.H., A.B., K.S.K., S.-B.T., A.T.L.T., L.W.J.T. are co-inventors of previously filed pending patents on artificial intelligence-based therapy development. The remaining authors declare no competing interests.

## Data availability

The data used in this study are available from the corresponding authors upon reasonable request.

## Code availability

Data processing and analyses in this study were conducted using basic functions written in Python programming language. The custom codes can be shared on reasonable request to the corresponding authors.

## Author information

These authors contributed equally: Shi-Bei Tan, Kirthika Senthil Kumar, Tiffany Rui Xuan Gan.

### Contributions

A.B., V.P.M., M.M.A., T.R.X.G., and D.H. conceived the study. V.P.M., M.M.A., T.R.X.G., D.H., A.B., S.-B.T., K.S.K., and A.T.L.T. designed the study. T.R.X.G. and V.P.M. extracted the data from medical records. S.-B.T., K.S.K., A.B., A.T.L.T., and L.W.J.T. analyzed the data. S.-B.T. and K.S.K. wrote early drafts. All authors contributed to scientific discussion, data interpretation, critical review, and approval of the final manuscript. S.-B.T., K.S.K., and T.R.X.G. are considered co-first authors.

## References

1. Cuenca, A.G., Kim, H.B. & Vakili, K. Pediatric liver transplantation. Seminars in Pediatric Surgery 26, 217–223 (2017).

2. Hackl, C., Schlitt, H.J., Melter, M., Knoppke, B. & Loss, M. Current developments in pediatric liver transplantation. World Journal of Hepatology 7, 1509 (2015).

3. McDiarmid, S.V. Management of the pediatric liver transplant patient. Liver Transplantation 7, S77–S86 (2001).

4. Woillard, J.-B., Saint-Marcoux, F., Debord, J. & Åsberg, A. Pharmacokinetic models to assist the prescriber in choosing the best tacrolimus dose. Pharmacological Research 130, 316–321 (2018).

5. Wallin, J.E., et al. Population pharmacokinetics of tacrolimus in pediatric liver transplantation: early posttransplantation clearance. Therapeutic drug monitoring 33, 663–672 (2011).

6. Taddeo, A., Prim, D., Bojescu, E.-D., Segura, J.-M. & Pfeifer, M.E. Point-of-Care Therapeutic Drug Monitoring for Precision Dosing of Immunosuppressive Drugs. The Journal of Applied Laboratory Medicine 5, 738–761 (2020).

7. Ng, V.L., et al. Barriers to ideal outcomes after pediatric liver transplantation. Pediatric transplantation 23, e13537 (2019).

8. Delaloye, J.-R., et al. Limited sampling strategies for monitoring tacrolimus in pediatric liver transplant recipients. Therapeutic drug monitoring 33, 380–386 (2011).

9. Kassir, N., et al. Population pharmacokinetics and Bayesian estimation of tacrolimus exposure in paediatric liver transplant recipients. British Journal of Clinical Pharmacology 77, 1051–1063 (2014).

10. Song, X., et al. Compare the performance of multiple machine learning models in predicting tacrolimus concentration for infant patients with living donor liver transplantation. Pediatric Transplantation, e14379 (2022).

11. Truong, A.T.L., et al. Harnessing CURATE.AI for N-of-1 Optimization Analysis of Combination Therapy in Hypertension Patients: A Retrospective Case Series. Advanced Therapeutics 4, 2100091 (2021).

12. Al-Shyoukh, I., et al. Systematic quantitative characterization of cellular responses induced by multiple signals. BMC Systems Biology 5, 88 (2011).

13. Blasiak, A., et al. IDentif. AI: Rapidly optimizing combination therapy design against severe Acute Respiratory Syndrome Coronavirus 2 (SARS-Cov-2) with digital drug development. Bioengineering & translational medicine 6, e10196 (2021).

14. Wang, H., et al. Mechanism-independent optimization of combinatorial nanodiamond and unmodified drug delivery using a phenotypically driven platform technology. ACS nano 9, 3332–3344 (2015).

15. Zarrinpar, A., et al. Individualizing liver transplant immunosuppression using a phenotypic personalized medicine platform. Science translational medicine 8, 333ra349–333ra349 (2016).

16. Zarrinpar, A., et al. Artificial intelligence based dosing of tacrolimus in liver transplantation: prospective, randomized Phase 2 trial. in 2022 American Transplant Congress, Vol. 106 129-129 (LIPPINCOTT WILLIAMS & WILKINS, 2022).

17. Abdulla, A., et al. Project IDentif.AI: Harnessing Artificial Intelligence to Rapidly Optimize Combination Therapy Development for Infectious Disease Intervention. Advanced Therapeutics 3, 2000034 (2020).

18. Blasiak, A., et al. The IDentif.AI-x pandemic readiness platform: Rapid prioritization of optimized COVID-19 combination therapy regimens. npj Digital Medicine 5, 83 (2022).

19. McCormack, P.L. & Keating, G.M. Tacrolimus. Drugs 66, 2269–2279 (2006).

20. Quak, S.H., Phua, K.B., Aw, M.M. & Krishnan, P. Liver transplantation in children: the Singapore experience. Singapore Medical Journal 62, S53 (2021).

