## Supplementary Material for "CURATE.AI – AI-derived personalized tacrolimus dosing for pediatric liver transplant: A retrospective study"

### Supplementary information

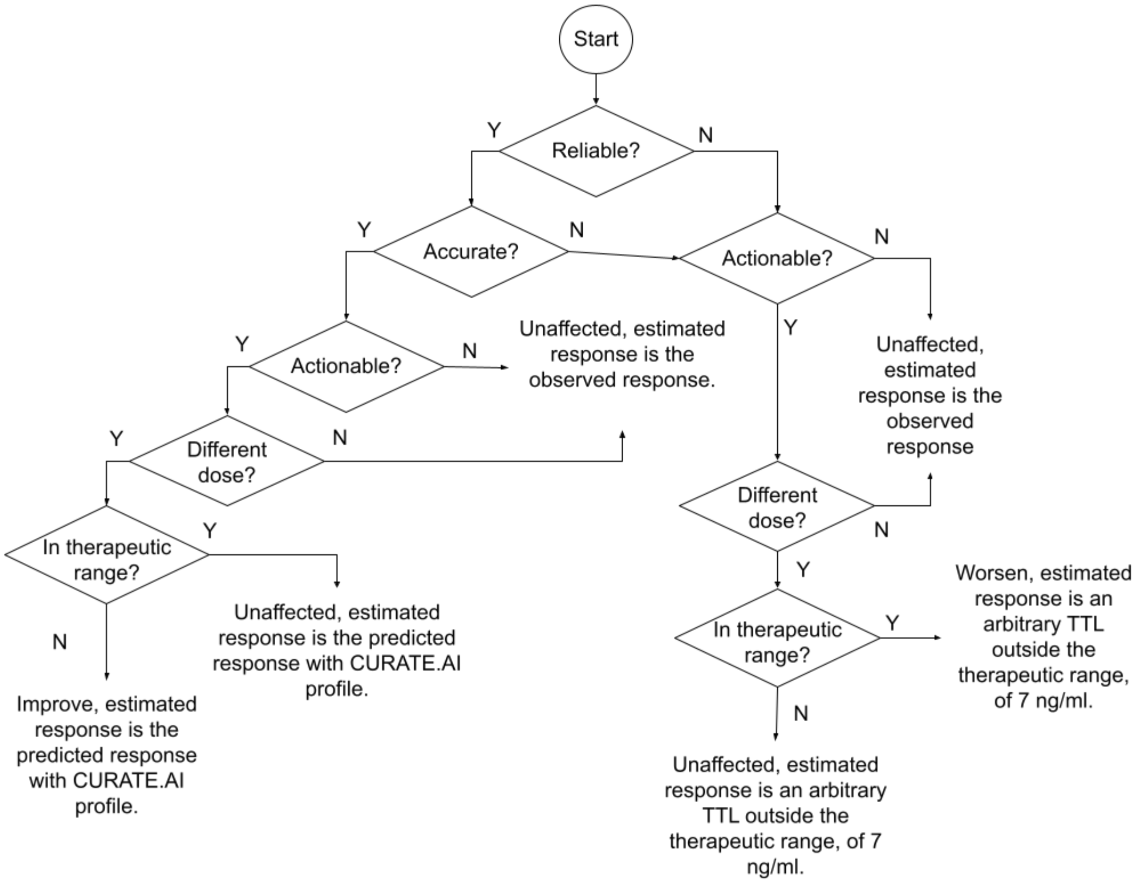


**Fig. S1 Criteria for assessing the effect of CURATE.AI on patient outcomes.** Reliable refers to the scenario where CURATE.AI correctly predicts within the therapeutic range or non-therapeutic range. Accurate refers to within clinically acceptable prediction error, pre-defined as between and inclusive of -1.5 and 2 ng/ml. Different dose refers to the different doses being recommended by CURATE.AI compared to the dose administered. Therapeutic range refers to scenarios within the therapeutic range, pre-defined as between and inclusive of 8 and 10 ng/ml. Actionable refers to scenarios where dose recommendations are 8 mg or below, as 8 mg is assessed to be a comfortable maximum dose. Improve means that the patient's TTL is expected to achieve the therapeutic range instead of the observed TTL which was outside the therapeutic range, and worsen means vice versa. Unaffected means that the TTL stays within the therapeutic range or outside the therapeutic range as per the observed TTL. Note that the arbitrary TTL set as 7 ng/ml was not used in any analysis in this study.

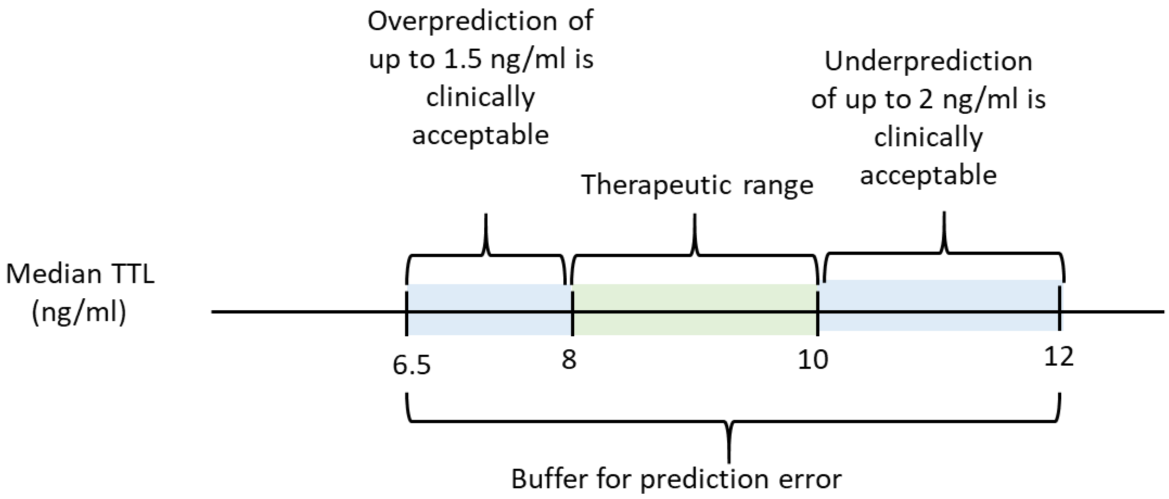


**Fig. S2: Clinically acceptable prediction error**. While the therapeutic range is between and inclusive of 8 and 10 ng/ml, 6.5 to 12 ng/ml was assessed as a clinically acceptable range. Thus, overprediction of up to 1.5 ng/ml and underprediction of up to 2 ng/ml are assessed to be clinically acceptable, and the clinically acceptable prediction error is between and inclusive of -1.5 to 2 ng/ml.
